# The DEXACELL Trial – a protocol for a pragmatic, multicentre, double-blind, placebo-controlled, randomised, parallel group, phase 3 superiority trial to assess the effectiveness and cost-effectiveness of DEXAmethasone as an adjunctive therapy for the management of CELLulitis in adults presenting to urgent secondary care in the UK

**DOI:** 10.1101/2025.08.26.25334247

**Authors:** Katherine Joyce, Rebecca Lear, Fergus Hamilton, David Arnold, Ella Chaudhuri, James Connors, Heather Cook, Siobhan Creanor, Phoebe Dawe, Elizabeth Goodwin, Annie Hawton, Chris Hayward, Daniel Lasserson, Matthew J Ridd, David Rowe, Debbie Shipley, Hazel Taylor, Hannah E Wainman, O. Martin Williams, Edd Carlton

## Abstract

**Introduction:** Cellulitis is a common bacterial skin infection causing significant pain, swelling, and impact on daily activities, frequently leading to emergency department presentations and hospital admissions. While antibiotics are the mainstay of treatment, they do not directly address inflammation, often resulting in persisting or worsening symptoms in the initial days. Corticosteroids, with their potent anti-inflammatory effects, have shown benefit in other acute infections but are not currently standard care for patients with cellulitis. This trial aims to determine if adjunctive oral dexamethasone can reduce pain and improve outcomes in adults with cellulitis presenting to UK urgent secondary care settings.

**Methods and analysis:** This is a pragmatic, multicentre, double-blind, placebo-controlled, randomised, parallel group, phase 3 superiority trial, with an internal pilot and parallel health economic evaluation. Adult patients (≥16 years) with a clinical diagnosis of cellulitis (at any body site except the orbit) presenting to urgent secondary care will be screened for eligibility. 450 participants will be randomised (1:1) to receive either two 8 mg doses of oral dexamethasone or matched placebo, administered approximately 24 hours apart, in addition to standard antibiotic therapy. The primary outcome is total pain experienced over the first 3 days post-randomisation, calculated using the standardised area under the curve (AUC) from pain scores (Numerical Rating Scale 0-10) across up to seven timepoints. Secondary outcomes include health-related quality of life (EQ-5D-5L), patient global impression of improvement, analgesia and antibiotic usage, hospital (re)admissions, complications, unscheduled healthcare use, cellulitis recurrence, and cost-effectiveness at 90 days. The primary estimand will apply a treatment policy approach to intercurrent events.

**Ethics and dissemination:** The trial has received ethical approval from South Central – Oxford B Research Ethics Committee (reference; 24/SC/0289) and will be conducted in compliance with GCP and applicable regulations. Informed consent will be obtained from all participants. Findings will be disseminated through peer-reviewed publications and conference presentations, and to patient groups and relevant clinical guideline committees.

**Trial registration:** ISRCTN76873478. Current protocol version 5.0, 7th January 2025.

**Strengths and limitations of this study:**

- This trial employs a robust double-blind, placebo-controlled design to minimise bias in assessing a subjective primary outcome (patient-reported pain).
- The pragmatic nature of the trial, recruiting from diverse urgent secondary care settings, with additional incentives to recruit from minoritised groups, aims to enhance the generalisability of findings to real-world clinical practice.
- Comprehensive follow-up to 90 days allows for assessment of both short-term symptom relief and longer-term impacts on healthcare utilisation and recurrence.
- The inclusion of a parallel health economic evaluation will provide crucial information on the cost-effectiveness of adjunctive dexamethasone.
- Potential variability in’usual care’ antibiotic regimens across sites, while reflecting real-world practice, is a possible limitation, which is accounted for in the pragmatic design.

## Introduction

### Background and rationale

Cellulitis, a common bacterial infection of the skin and subcutaneous tissues, imposes a substantial burden on both patients and healthcare systems.^1^ It is characterised by pain, swelling, erythema, and warmth, often leading to reduced mobility and ability to perform daily activities. In England alone, cellulitis accounts for over 300,000 presentations to Emergency Departments (EDs) annually, with approximately 50% of these patients requiring hospital admission.^2^ Patients with cellulitis represent about 3% of all adult hospital admissions and are estimated to occupy 1% of NHS hospital beds in England and Wales.^3^

Standard UK management of cellulitis involves antibiotic therapy, analgesia, and elevation of the affected limb.^4^ Despite antibiotic treatment aimed at eradicating the bacterial cause, the associated inflammation can persist or even worsen in the initial 48-72 hours.^5^ This ongoing inflammation often manifests as significant pain, which is a major reason for patient re-attendance at hospitals or other healthcare providers, occurring in approximately one in five patients.^6,7^ Such re-presentations can lead to extended or alternative antibiotic courses, which may offer no additional benefit whilst contributing to increased costs and antibiotic resistance.^8,9^ There is therefore interest in improving early symptomatic response in cellulitis by modulating the host inflammatory response.

Oral corticosteroids are well-established anti-inflammatory agents shown to be effective in numerous other acute infectious and inflammatory conditions to dampen the immune response and improve short-term symptoms. For example, systematic reviews have shown benefits of adjunctive corticosteroids in conditions like sore throat and in croup.^10,11^ Two previous randomised trials have investigated corticosteroids for acute cellulitis. One Danish trial (n=112) found that prednisolone (30 mg daily, reducing over a week) significantly reduced time to clinical cure compared to placebo (10.0 vs 14.6 days, p<0.01), with no evidence of increased recurrence of cellulitis at 1-year follow-up.^12,13^ An unpublished trial (NCT01671423, n=25) reported a non-statistically significant trend towards greater pain reduction at 48 hours with a single dose of 60 mg prednisolone versus placebo (mean change from baseline 39.9 vs 30.5 points on a 0-100 visual analogue scale (VAS)). Additionally, trials of non-steroidal anti-inflammatory drugs (NSAIDs), which may have a similar mechanism of action to corticosteroids, have suggested a benefit in cellulitis.^14^ However, there are concerns about the adverse effects of NSAIDs and so these were not considered in this trial. Current guidelines from the Infectious Diseases Society of America (IDSA), acknowledge the potential for corticosteroids for the treatment of cellulitis but call for further high-quality randomised controlled trials (RCTs) to establish the role of corticosteroids in cellulitis management, a position also echoed by Cochrane.^15,16^

The DEXACELL trial is designed to address this evidence gap. It is a pragmatic, multicentre, double-blind, placebo-controlled, randomised, parallel group, phase 3 superiority trial with an internal pilot phase and a parallel health economic evaluation.

The primary research question is: *Is the addition of oral dexamethasone to usual care in patients who present to urgent secondary care with cellulitis effective and cost-effective in terms of reducing pain, improving quality of life, and reducing further antimicrobial usage and healthcare utilisation?* Given the high incidence of cellulitis, even a modest improvement in symptoms and/or a reduction in healthcare costs could have significant population-level benefits.

### Objectives

*Primary objective:* To establish if the addition of dexamethasone to treat patients presenting to urgent secondary care with cellulitis reduces total pain reported over the first three days (post-randomisation) compared to a control (placebo).

*Secondary objectives:* To determine whether the addition of dexamethasone, when compared to a control (placebo), to treat patients with cellulitis presenting to urgent secondary care with cellulitis: a) improves quality of life and other patient-reported outcomes, b) reduces subsequent antimicrobial prescribing, analgesia usage and healthcare utilisation and c) is cost-effective.

## Methods and analysis

### Trial design and setting

This is a pragmatic, multicentre, double-blind, placebo-controlled, randomised, parallel group, phase 3 superiority trial with an internal pilot and parallel health economic evaluation. Potential participants will be identified and recruited from urgent secondary care services (e.g. Emergency Departments, Ambulatory Care Units, Same Day Emergency Care) across up to 20 sites in the UK. After providing informed consent, participants will be individually randomised on a 1:1 ratio to receive either oral dexamethasone or matched placebo in addition to standard antibiotic therapy.

### Eligibility criteria

The eligibility criteria for the trial are detailed in Table 1, below.

**Table 1:** Inclusion and exclusion criteria.

| Inclusion criteria | Exclusion criteria |
| --- | --- |
| *Participants must meet all of the following:* | *Participants must meet none of the following:* |
| Aged 16 years old or over.<br>A current clinical diagnosis of cellulitis at any body site except the orbit (periorbital/orbital cellulitis).<br>Able to provide informed consent. | Orbital or periorbital cellulitis, surgical site infection, or planned surgical management (e.g., abscess) as managed under a different clinical pathway.<br>Known allergy to dexamethasone.<br>Contraindication to dexamethasone due to concurrent medication (e.g., cobicistat).<br>Known current invasive fungal infection<sup>1</sup>.<br>Known current gastric or duodenal ulceration.<br>Already on systemic corticosteroids (other concomitant corticosteroids such as inhalers or creams are acceptable).<br>Unable to take oral medication.<br>Lack of capacity.<br>Inability to complete follow-up procedures. |
| *People of child-bearing potential must be willing to:* |  |
| Use a highly effective method of contraception (and must agree to continue 3 months after the last dose of the IMP).<br>Inform the trial team if pregnancy occurs during trial participation. |  |
|  | Prisoner<sup>2</sup>. |
|  | *People of child-bearing potential only:* |
|  | Pregnant, breastfeeding, or planning to conceive in next 3 months. |

<sup>1</sup>This includes only invasive infections such as pulmonary aspergillosis and does NOT include cutaneous infections such as athlete's foot, vaginal thrush, etc.
<sup>2</sup>This does not exclude patients in police custody, though consideration should be given to whether they are able to complete the trial follow-up procedures.

### Intervention and comparator

#### Intervention

Participants in the intervention group will receive two 8 mg doses of oral dexamethasone. The first dose will be administered as soon as possible after randomisation, whilst in the urgent secondary care unit. The second dose is to be taken 24 hours later (±6 hours). If discharged, participants will take the second dose at home; if hospitalised, it will be administered by clinical or research staff. Each 8 mg dose consists of two 4 mg dexamethasone tablets over-encapsulated into two separate capsules for blinding purposes. No modification of trial dosage is permitted in the study.

Dexamethasone was chosen for its high glucocorticoid activity and recent clinical experience in trials like RECOVERY.^17^ The dose (8 mg), route (oral), and course (2 doses) were based on expert opinion suggesting additional benefit of a second dose at ∼24 hours for pain outcomes. Given its half-life, the selected regimen is expected to affect swelling and pain over the subsequent 24-48 hours.

#### Comparator

Participants in the comparator group will receive two doses of matched placebo capsules, identical in appearance and administered on the same schedule as the active drug. The placebo design is crucial to ensure blinding due to the participant reported primary outcome.

#### Usual care

All participants will receive usual care for cellulitis as per local policy at each site, including; clinical assessment, antibiotics and analgesia, hospital admission if required, advice on management of concomitant conditions and expected symptom duration. Details of any antibiotics and analgesia administered will be recorded at baseline and at day 14 follow-up.

#### Compliance

Intervention compliance for dose 1 and any reason for non-compliance will be recorded on the study database by site staff. Intervention compliance for dose 2 will be self-reported by participants via yes/no response to a SMS message. Compliance rates will be monitored by the oversight committees.

### Outcomes

#### Primary outcome

Total pain experienced over the first 3 days post-randomisation.

Total pain will be calculated using the standardised area under the curve (AUC) approach from seven individual pain scores, measured using a 0-10 numerical rating scale (NRS) (see wording in Appendix 2). The first (baseline) pain score is collected in person pre-randomisation as part of a participant questionnaire pack. The remaining six scores are collected post-randomisation at approximately 12-hour intervals (around 8 am and 8 pm) via SMS survey (hereby referred to as timepoints 1-6 (T1-T6)). Actual time of response will be recorded. Alternative methods to collect pain scores (in-person, telephone) are available if SMS is not feasible, depending on staff capacity.

Total pain was chosen as the primary outcome after extensive Patient and Public Involvement (PPI) work. Reduction in erythema has previously been used as a cellulitis outcome measure by regulators^18^ but was not chosen in this trial for multiple reasons. Firstly, patient feedback was that pain and symptoms were much more important than the size of the erythema. Secondly, measurement of erythema might be biasing against those of skin of colour. Pain was identified as a key issue for patients with cellulitis, aligning with recent priority setting partnerships.^19^

#### Secondary outcomes

For the secondary outcomes, listed below, continuous outcomes will be reported as means and binary / categorical outcomes will be reported as percentages at the specified timepoints.

1. Health-related quality of life, measured by EQ-5D-5L^20^ at Day 3, Day 14 and Day 90 post-randomisation.
2. Patient Global Impression of Improvement (PGI-I^21^) measured daily for first 3 days (via SMS at T2, T4, T6) and at Day 14 post-randomisation.
3. Analgesia usage (number and type of analgesia taken over first 3 days) post-randomisation, collected at Day 14.
4. Antibiotic usage (route, type, and post-randomisation length of course) up to Day 14 post-randomisation.
5. (Re)admissions to hospital by Day 14 post-randomisation.
6. Complications of dexamethasone use by Day 14 post-randomisation.
7. Unscheduled healthcare usage until Day 14 post-randomisation.
8. Health, social care and broader societal resource use, measured by a resource use questionnaire based on the Modular Resource Use core module (ModRUM^22^) tailored to the study population, to Day 90 post-randomisation.
9. Recurrence of cellulitis by Day 90 post-randomisation.
10. Serious and/or potentially related adverse events by Day 90 post-randomisation.
11. Pain experienced at Day 14 post-randomisation, measured using NRS^23^ (0-10).

### Data collection methods

Data will be collected using electronic Case Report Forms (eCRFs) within a validated REDCap (Research Electronic Data Capture) Academic system provided by the UKCRC-registered Exeter Clinical Trials Unit (ExeCTU). All staff delegated to collect data in this trial will be appropriately trained.

Baseline data will be collected by site staff in person, with the exception of participant-reported outcome questionnaires (EQ-5D-5L and resource use questionnaire) which are self-completed by the participant. Minimum baseline data which must be collected prior to randomisation include: eligibility and consent data, baseline pain score, mobile phone number, and the minimisation factors (diabetes status, severity of cellulitis, prior antimicrobial therapy for current episode of cellulitis). Remaining baseline data will be collected ideally before randomisation, or after randomisation but prior to administering the first dose of the allocated treatment. This is to allow flexibility for swift recruitment in the emergency setting.

Subsequent pain scores (T1-T6) and PGI-I (T2, T4, T6) are completed electronically by the participant via SMS, or by site staff directly onto the trial REDCap system if conducting follow-up in-person/telephone.

Day 14 and Day 90 data are recorded by site staff during follow-up telephone calls. Participants receive an automated SMS message to remind them of the appointment, and research staff may try contacting the participant up to three times in order to promote complete follow-up and minimise missing data. If a participant is not engaging with the study at a particular follow-up timepoint, attempts will still be made to contact them at the next timepoint.

PeRSEVERE principles will be followed for participants who cease to engage with the study (See: https://persevereprinciples.org/). Unless a participant expressly indicates they wish to fully withdraw from the trial, attempts will be made to collect all follow-up data from randomised participants regardless of intervention compliance; if a participant becomes uncontactable and stops engaging with the study, passive data collection will continue where possible and available from the participant’s routine medical notes (e.g. (re)admissions at Day 14).

### Potential harms

Adverse events will be reviewed with participants at the Day 14 and Day 90 follow-up timepoints. All adverse events occurring from randomisation up to 90-days after that are either deemed definitely, probably or possibly related to the intervention (adverse reaction; AR) or meeting the definition of seriousness as detailed below (serious adverse event; SAE) or both (serious adverse reaction; SAR) will be recorded in the study database and monitored by oversight committees.

All SAEs and SARs will be reported to ExeCTU within 24 hours of the site becoming aware for onward reporting to the Sponsor. The trial chief investigator (or delegate) will assess the relatedness of all reported SAE/SARs, and if deemed related will assess the expectedness using the dexamethasone summary of product characteristics as the reference safety information. If an event is deemed related and unexpected (suspected unexpected serious adverse reaction; SUSAR) the Sponsor will onward report to the MHRA as required.

Serious adverse event is any untoward medical occurrence that:

- Results in death.
- Is life-threatening; an event in which the participant was at risk of death at the time of the event. This does not refer to an event which hypothetically might have caused death if it were more severe.
- Requires inpatient hospitalisation or prolongation of existing hospitalisation.
- Results in persistent or significant disability/incapacity.
- Consists of a congenital anomaly or birth defect.
- Other ‘important medical events’ may also be considered serious if they jeopardise the participant or require an intervention to prevent one of the above consequences.

The dosage regimen in this trial is a relatively low dose for a short period, meaning the risk of harm is low. In this trial the following conditions should always be reported as serious adverse events, to ensure adequate monitoring, due to the risk of them occurring as side effects of the study drug in our trial population:

- Severe hyperglycaemia (ketoacidosis, hyperglycaemic hyperosmolar state or hyperglycaemia requiring new use of insulin)
- Gastrointestinal bleeds
- Psychosis

The above risks are identified as associated with short-term use of corticosteroids. Further details of how these risks will be managed in the trial can be found in Appendix 1.

### Participant timeline

All screening, eligibility, consent and baseline data will be collected in-person during attendance to secondary urgent care. The participant will be asked to complete a short baseline questionnaire pack, and the study team will collect the remaining baseline data (see Table 2). Once the minimum baseline data is collected (see data collection section); participants will be randomised and administered the first dose of their allocated intervention. Early follow-up (T1-T6) occurs via SMS survey approximately every 12 hours for the first 3 days, starting at the next available 8am or 8pm timepoint after randomisation. This includes the collection of pain scores to calculate the primary outcome plus some minimal data for secondary outcomes. Additional follow-up to collect secondary outcome data via telephone occurs at Day 14 (± 2 days) and Day 90 (± 7 days). Participants will be advised to use a paper diary/aide-memoire to record data to help them during the Day 14 and Day 90 follow-up calls.

**Table 2:**
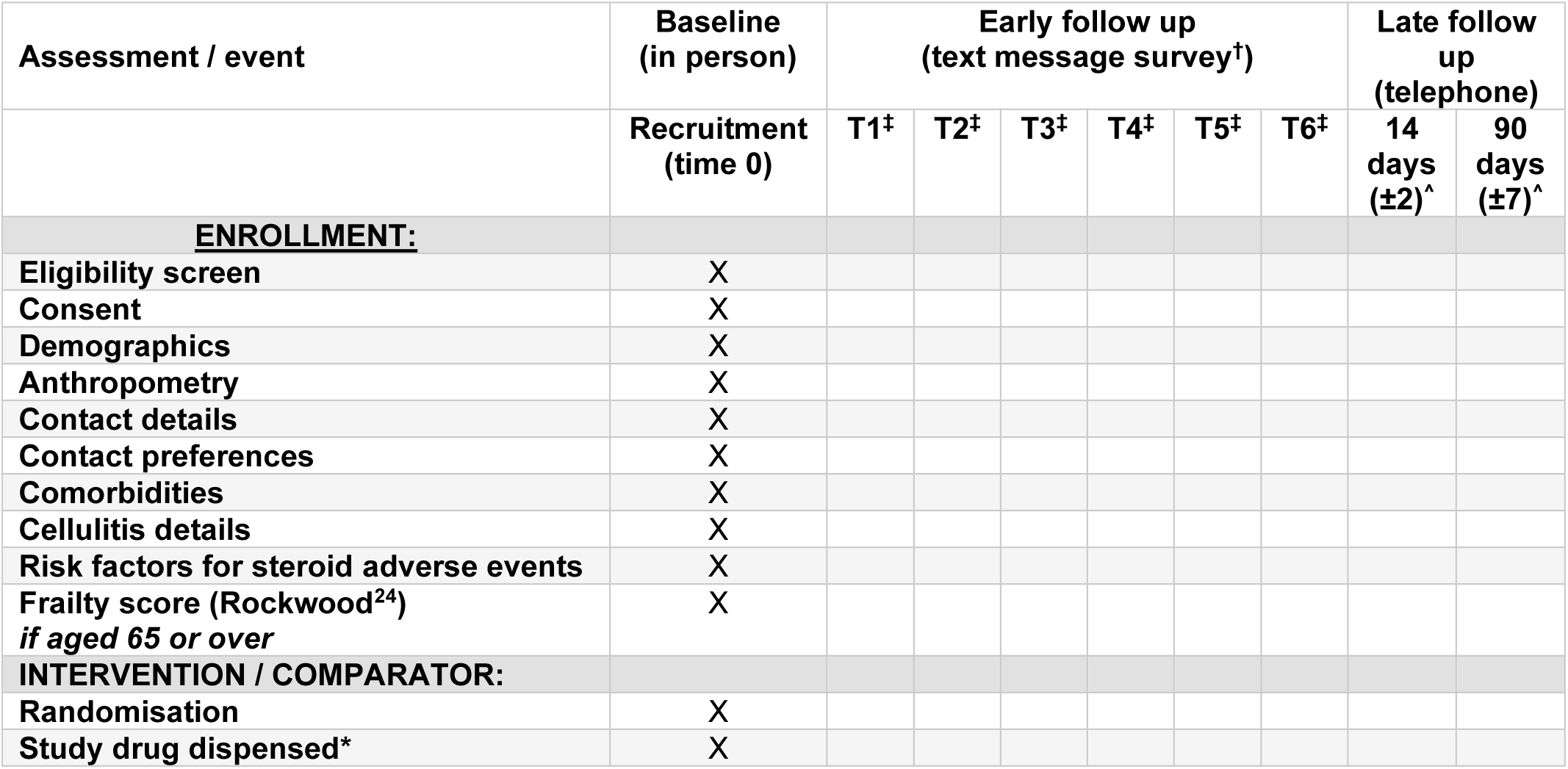
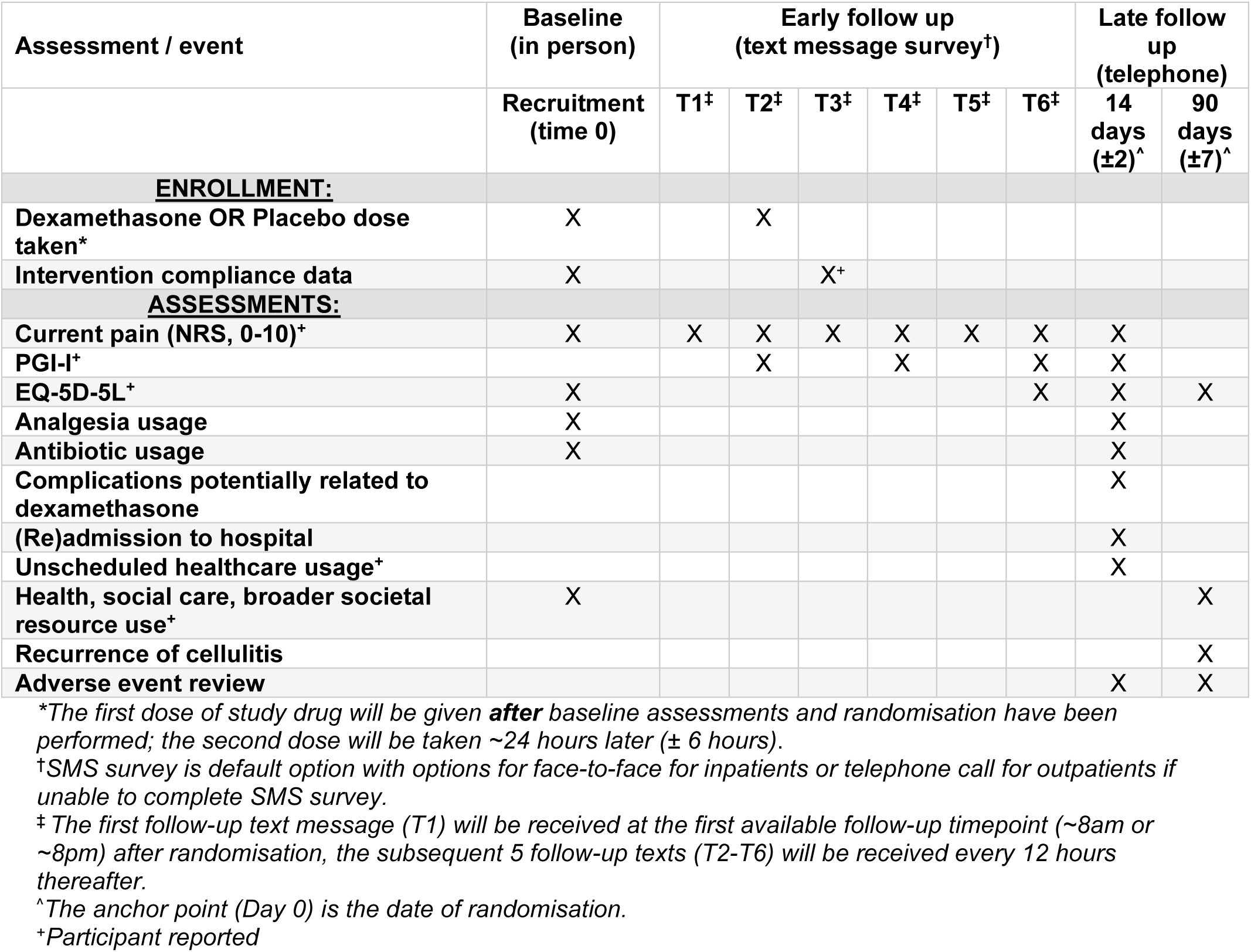
Data collection and schedule of assessments.

### Sample size

The sample size was calculated to detect a minimum clinically important difference of 10 points^25^ (on a 0-100 standardised AUC scale for pain) between allocated groups over the first 3 days post-randomisation.

Assuming a standard deviation (SD) of 30 points (a conservative estimate based on previous cellulitis trials and meta-analyses^12,14,26^), 191 participants per group with primary outcome data are needed for 90% power at a two-sided 5% statistical significance level. To account for up to 15% of participants not returning sufficient pain scores to derive the primary outcome (baseline pain score plus at least one more pain score at least 24 hours later), the recruitment target is 450 participants (225 per group). The sample size was calculated using PASS sample size software. The SD estimate will be reviewed, blinded, at the end of the internal pilot phase.

### Recruitment

Potential participants will be identified and screened by local research teams or clinicians on presentation to a participating urgent secondary care site. This will be completed via review of medical records and discussion with the potential participant, as needed. If potentially eligible they will be invited to review the participant information materials and, if they are interested, to provide their informed consent to participate.

Final eligibility will be confirmed by the local site principal investigator or an appropriately qualified and delegated clinician (registered prescriber with appropriate clinical experience). If the potential participant is of child-bearing potential, they will be required to complete a pregnancy test before final eligibility can be confirmed.

Efforts to ensure inclusive recruitment include; selecting participating sites to cover underserved geographical areas, recording any language barriers during the internal pilot phase to identify needs for translation of key documents (PIS, video subtitles), and specific funding for sites to support recruitment of people who inject drugs, an underserved population with high rates of skin infection.

Efforts have been made to minimise staff and participant burden to encourage recruitment in an urgent secondary care setting by minimising data collection as far as possible and designing all follow-up to be remote, negating the need for additional in-person visits for the trial.

### Randomisation: sequence generation, allocation concealment mechanism, and implementation

Randomisation will be stratified by recruiting site and then minimised using an algorithm with a random element to balance allocated groups on three factors:

1. Prior antimicrobial therapy for this current episode of cellulitis (yes/no)
2. Diabetes status (defined by a known diagnosis of either type 1 or type 2 diabetes mellitus, yes/no)
3. Severity of Cellulitis (Eron Class 1 vs all other classes).

Eligible, consented participants with minimum baseline data collected will be randomised 1:1 to either dexamethasone or placebo. Randomisation is carried out via a secure 24-hour web-based randomisation service provided by the Centre for Healthcare Randomised Trials (CHaRT, University of Aberdeen).

Randomisation will be completed by a delegated member of the site team or (in case of technical issues at site) may be conducted by ExeCTU staff on direction from a delegated member of the site team.

A master list of pack IDs will be produced by the senior (unblinded) statistician and provided to CHaRT for upload into the randomisation system and to the IMP manufacturer for labelling of packs. The master list will indicate which packs contain placebo and which contain dexamethasone.

The CHaRT system will be integrated into the main study database (REDCap) so that randomisation can be initiated from within REDCap. Once the online randomisation process is complete, REDCap will indicate to the user a blinded pack ID which should be dispensed to the participant; it will not indicate whether the participant has been allocated to receive IMP or placebo. All site staff (and any central trial staff who may assist sites with randomisation) will be blinded to allocation and will not have any access to the master list of pack IDs or allocation sequence.

### Blinding

The trial is double-blind; participants and site research teams (clinicians, data collectors, outcome assessors) will be unaware of treatment allocation. This is achieved by the IMP manufacturer providing identically appearing over-encapsulated IMP (each containing a 4 mg dexamethasone tablet with backfill) and matching placebo capsules, packaged and labelled with blinded pack IDs. Inability to take oral medication is listed as an exclusion criterion to prevent participants or staff becoming accidentally unblinded by opening the capsules to administer the intervention.

Access to the master list linking pack IDs to treatment allocation will be restricted to the senior (unblinded) statistician, the IMP manufacturer, developers of the randomisation system (CHaRT), and unblinded members of ExeCTU team as required for their role. The randomisation system will be fully tested prior to go-live to ensure the adequate concealment of allocation. Analysing trial statisticians will remain blinded until the primary statistical analysis of the primary outcome is complete.

Emergency unblinding will be available 24/7 via a dedicated automated phoneline in case clinically necessary for the medical management of a participant. The decision to unblind rests with the treating clinician. Unblinding events will be documented and reported.

### Data management

ExeCTU will oversee the day-to-day management of participant data according to a trial specific Data Management Plan (DMP). Work instructions and ongoing training will be provided to recruiting site teams on record keeping and data entry processes.

All screening, baseline and outcome data for the trial will be entered into an electronic data capture system (EDC; REDCap Academic), hosted by University of Exeter and managed by ExeCTU following UK General Data Protection Regulation. Trial specific eCRFs will be created including validation and range checks at the point of data entry. Data will be entered into the eCRFs via a combination of SMS and online surveys to participants, and entry into the database by site staff. Full testing of the EDC will be conducted and documented before starting recruitment.

Access to REDCap Academic for the trial will be managed using password protected individual user accounts. Each account will be restricted by site, functionality and data as appropriate for the person’s role. REDCap Academic also maintains electronic audit trails to ensure data integrity and security. Access will be granted to authorised representatives from the North Bristol NHS Trust as Sponsor, as well as representatives from University of Exeter and regulatory agencies e.g. MHRA, if required for the purposes of auditing, monitoring and inspection of the trial.

The essential documentation will be archived for a minimum of 10 years after the end of the trial. After 10 years, all personal identifiable data will be securely destroyed upon authorisation from the Sponsor. The anonymised dataset will be stored indefinitely for the purposes of future ethically approved research.

## Statistical methods

A detailed Statistical Analysis Plan (SAP) will be written following Gamble et al^27^ guidelines, approved by an independent statistician, and made publicly available before the trial follow-up period concludes. Any amendments to the SAP will be documented. Results will be reported following the CONSORT statement and relevant extensions (e.g., for Patient Reported Outcomes, Pragmatic Trials). Emphasis will be on estimation rather than hypothesis testing; tests will be at the 5% two-sided level. Baseline characteristics will be summarised descriptively overall and by allocated group (means/SDs or medians/interquartile ranges for continuous; Ns/% for categorical), with no formal between-group testing of baseline data. A CONSORT flow diagram will illustrate participant flow.

The primary analysis population will be all randomised participants who contribute at least 2 out of a possible 7 pain scores, with the second pain score being at least 24 hours after the baseline pain score. The primary outcome (pain AUC, standardised 0-100) will be analysed using a mixed-effects linear regression model, adjusting for baseline pain, age, sex, and the minimisation variables (severity of cellulitis (Eron Stage^28^ 1 vs all other stages), antimicrobial therapy for this episode prior to attending urgent secondary care, diabetes status) as fixed effects, and recruitment site as a random effect to account for potential between-site heterogeneity. The adjusted difference in means (and 95% CI) between dexamethasone and placebo groups will be reported; unadjusted difference will also be reported. The primary estimand addresses the effect of dexamethasone plus usual care vs. placebo plus usual care, with a treatment policy strategy for intercurrent events. Missing pain scores for the primary outcome will be imputed using linear interpolation if scores are available on either side; if final scores are missing, last observation carried forward (a conservative approach) will be used. Sensitivity analyses will explore other imputation methods (e.g., multiple imputation using chained equations including allocated group, baseline score, centre, minimisation variables, age, sex, and predictors of missingness). If an imbalance in baseline characteristics thought to be predictive of outcome is noted, the analysis will be repeated adjusting for these.

Secondary continuous outcomes (PGI-I, pain at 14 days) will be analysed using mixed-effects linear regression models if assumptions met; PGI-I at days 1, 2, and 3 will use a mixed-effects repeated measures model. Alternative modelling (e.g., mixed-effects ordinal logistic models, dichotomisation for logistic regression) will be considered if assumptions not met, detailed in SAP. Dichotomous secondary outcomes (additional antibiotic use, (re)admission, unscheduled healthcare use, recurrence) will be analysed using mixed-effects logistic models (if event numbers allow). All models will adjust for the same factors as the primary analysis. Adjusted differences in means or odds ratios (with 95% CIs) will be reported, along with unadjusted estimates.

Descriptive analysis will cover compliance, analgesia details, antibiotic details, and dexamethasone-related complications. The estimands framework for secondary outcomes will mirror the primary outcome. Multiple imputation may be considered for missing secondary outcome data. SAEs will be reported descriptively by whether the first dose was taken (not ITT).

Exploratory subgroup analyses on the primary outcome (not powered for) will be pre-specified for: cellulitis location (lower limb vs other) and NSAID usage at randomisation (user vs non-user). The primary analysis model will be refitted including an interaction term between treatment and the subgroup of interest; minimum numbers for these analyses will be pre-specified in the SAP.

### Cost-effectiveness analysis

A full within-trial cost-effectiveness analysis (CEA) will be conducted from a NHS and Personal Social Services perspective to estimate the incremental cost-effectiveness of dexamethasone versus placebo.

Resources and costs of providing oral dexamethasone will be established. Participant health, social care, and broader societal resource use will be captured at baseline and Day 90 using a self-report resource use questionnaire tailored for this population with PAG input. Nationally-recognised UK health and social care unit costs will be applied. Quality-adjusted life-years (QALYs) will be estimated using EQ-5D-5L data collected at baseline, Day 14, and Day 90. The’cross-walk’ algorithm will map EQ-5D-5L to EQ-5D-3L UK general population valuation survey data, per NICE guidance. Descriptive statistics will summarise costs and QALYs by group. Mixed-effects linear regression models will test for differences in costs and QALYs, adjusting for age, sex, baseline pain, severity of cellulitis (Eron Stage^28^ 1 vs all other stages), prior antimicrobial therapy for this episode prior to attending urgent secondary care, diabetes status (fixed effects), and recruitment site (random effect); cost models will also adjust for baseline costs, and QALY models for baseline EQ-5D-5L values. ICERs will be presented for cost per total pain prevented (3 days) and cost per QALY (Day 90). Sampling uncertainty will be accounted for, missing data explored for multiple imputation, and cost-effectiveness acceptability curves presented using the net-benefit approach against NICE thresholds (e.g., £20,000-£30,000 per QALY). Broader societal perspectives will be considered in sensitivity analyses. A Health Economics Data Analysis Plan concordant with the trial SAP and CHEERS^29^ guidelines will be developed.

### Patient and public involvement

A patient advisory group (PAG) will advise on study processes throughout the trial and includes the Patient and Public Involvement (PPI) co-applicant, North Bristol NHS Trust infection PAG and three additional members who have relevant lived experience and have been identified to join the group specifically for DEXACELL. The PAG will be involved at every stage of the trial including but not limited to; reviewing and inputting into patient-facing materials throughout the trial, advising on the participant pathway and any potential recruitment barriers, and co-producing the dissemination materials to be shared with participants and the public once the trial results are available.

The study team have also worked with people who inject drugs who commonly suffer with cellulitis, via the Bristol Drugs Project, and patients with skin of colour where cellulitis looks different. This has helped to design an inclusive trial to ensure our research is relevant to these important groups.

The co-applicant patient representative sits on the Trial Management Group (TMG) and two independent lay members sit on the Trial Steering Committee (TSC).

### Trial oversight committees

An independent Data Monitoring Committee (DMC) will meet remotely approximately every 6-months to review reports of accumulating, pooled and unblinded trial data, to monitor the progress and conduct of the trial, safeguard the interests of trial participants and assess the safety and efficacy of the interventions during the trial. This will include unblinded data. The members of the DMC for this trial are Prof Beth Stuart (Chair, Statistician, Queen Mary University of London), Dr James Foley (Clinician, University Hospital Galway) and Dr Rebecca Sutherland (Clinician, NHS Lothian). DMC members are independent of the trial team, Sponsor and Funder. The senior trial statistician will be unblinded and prepare/review unblinded DMC reports. The DMC will report their recommendations to the independent Trial Steering Committee (TSC). The TSC will review the final protocol, oversee trial progress, review the SAP, and make the final decision on early trial termination if recommended by the DMC. Both the DMC and TSC will work to charters agreed with ExeCTU, available on request.

No formal interim analyses for effectiveness or futility are planned. The DMC and TSC will review accumulating trial data at the end of an internal pilot phase (after the first 6 months of participant recruitment) to assess feasibility of the trial. This phase will focus on targets to open ≥10 sites, recruit ≥70 participants, achieve a mean recruitment rate of ≥2 recruits/site/month, and ≥85% of participants with calculable primary outcome. If all of these criteria are met the trial will proceed. If not, progression criteria will be assessed on a Green/Amber/Red traffic light system (detailed in full protocol) which will guide trial continuation in consultation with the DMC, TSC, Sponsor and funder.

### Trial monitoring

Central monitoring will be completed by ExeCTU and includes; review of delegation and training logs of site staff to ensure appropriate training and delegation of staff confirming eligibility, consenting, randomising, and dispensing IMP, and monitoring of consent forms to ensure completeness and adherence with Good Clinical Practice (GCP). The data management team will conduct regular data cleaning checks and provide reports to the TMG detailing key data and its completeness throughout the trial.

Remote monitoring is planned for once per site after at least 5 participants have been recruited, or after 6 months of the site opening, and includes the review of prescriptions, IMP temperature logs and IMP accountability logs, and review of the medical notes to ensure adequate documentation of participation and eligibility. Extra on-site monitoring may be conducted if triggered, or if concerns are raised by an individual with knowledge of the trial.

Further details of monitoring and data validation activities are outlined in a monitoring plan and DMP, available upon request.

## Discussion

The DEXACELL trial is designed to provide robust evidence on the clinical and cost-effectiveness of adjunctive oral dexamethasone for adults presenting with cellulitis to urgent secondary care. Cellulitis remains a significant cause of patient morbidity and healthcare resource utilisation.^1,2,30^ While antibiotics target the bacterial infection, the inflammatory component, which drives pain and systemic symptoms, is often inadequately addressed in the crucial early phase of illness. This trial hypothesises that a short course of dexamethasone, a potent anti-inflammatory agent, can lead to more rapid symptom resolution, particularly pain reduction, thereby improving patient experience and potentially reducing downstream healthcare use, such as re-consultations and hospital admissions.

Extensive Patient and Public Involvement (PPI) work during the development of this trial identified pain as a key issue in patients with cellulitis, aligning with recent priority setting partnerships, and participant-reported pain was therefore chosen as our primary outcome. The choice of a pragmatic, placebo-controlled, double-blind design is a key strength, minimising bias in the assessment of this subjective primary outcome which would be at great risk of bias in an open-label trial design and enhancing the generalisability of the findings to typical UK urgent secondary care settings. The inclusion of a diverse range of secondary outcomes, including quality of life, patient-reported improvement, and healthcare utilisation, will provide a comprehensive assessment of the impact of dexamethasone. The integrated cost-effectiveness analysis is vital for informing healthcare policy and resource allocation decisions, should dexamethasone prove effective.

Potential challenges include achieving the recruitment target within the planned timeframe, particularly given the seasonal variation in cellulitis incidence.^2^ The internal pilot phase is designed to assess and mitigate these risks early on. Another ongoing trial, COAT,^31^ is recruiting participants with cellulitis, comparing the efficacy and safety of 5 days versus 7 days of antibiotic treatment. However, this trial is not anticipated to impact recruitment for DEXACELL as they are primarily recruiting in primary care settings.

Ensuring adherence to the second dose of medication, taken at home by many participants, will also be important, though the short two-dose regimen is expected to facilitate compliance.

The chosen dose and duration of dexamethasone (total 16 mg over 24 hours) are relatively low compared to its use in other conditions and is much lower than doses associated with severe adverse effects in other contexts; extensive experience suggests a favourable safety profile for short courses in infection. However, the trial incorporates measures to manage potential risks associated with short-term corticosteroid use (e.g., hyperglycaemia in patients with diabetes and gastrointestinal upset), such as eligibility screening, site training and resources, additional patient information, and monitoring (further details are available in Appendix 1).

The active involvement of patients and the public (PPI) throughout the trial’s design (e.g., input on outcomes, patient-facing materials, data collection methods) and planned conduct, including specific engagement with underserved groups like people who inject drugs and people with skin of colour, strengthens its relevance and inclusivity.

If this trial demonstrates that adjunctive dexamethasone is effective and safe for treating cellulitis in urgent secondary care settings, it has the potential to change clinical practice for a very common and burdensome condition. This could lead to improved patient outcomes, such as faster pain relief and recovery, and more efficient use of healthcare resources by potentially reducing re-consultations, antibiotic courses, and hospital admissions. The findings will be disseminated widely through academic publications, presentations, and to patient and public groups and guideline committees to ensure they reach clinicians, patients, and policymakers, thereby facilitating translation into practice.

## Ethics and dissemination

### Research ethics approval

The trial protocol (version 5.0, 7th January 2025) and associated documents have been approved by an NHS Research Ethics Committee (IRAS number: 1009877) and have regard for HRA guidance.

### Dissemination policy

The trial is planned to take three years to complete from grant opening, with dissemination of results planned for late 2026. The trial website and social media accounts will share updates as the trial progresses. Trial findings will be disseminated through usual academic channels including peer-reviewed publications, presentations at international meetings, through patient organisations and participant newsletters (where consent is given). Patient groups will guide the development of patient-facing dissemination of results.

Results will also be communicated to the Royal College of Emergency Medicine, Society for Acute Medicine, NICE, and NHS England to inform national guidelines. The results will also be posted on the ISRCTN registry and trial website.

Authorship on publications will follow the International Committee of Medical Journal Editors (ICMJE) guidance.

### Protocol amendments

Any amendments to trial documents will be categorised and approved by the trial Sponsor before submission to HRA/REC/MHRA, as required. Substantial and relevant non-substantial amendments will be discussed by the TMG, PAG and TSC as appropriate. The funder representative will be notified of all amendments to the protocol and approved amendments are communicated by ExeCTU to participating sites for implementation. The chief investigator (or delegate) will inform the trial registry of any amendments.

### Consent

Written informed consent will be obtained from all participants by an appropriately trained and delegated member of the research team at the participating site prior to any trial-specific procedures, including randomisation and pregnancy testing (if not routine practice at site). Consent will be sought face-to-face, using either a paper or electronic consent form, depending on site preference. The Principal Investigator at each site is responsible for ensuring that any staff they delegate to receive informed consent is appropriately trained; this must include Good Clinical Practice training.

If a participant has capacity to consent but cannot physically sign the consent form (e.g. due to cellulitis in the hand), a witness independent of the trial may sign the consent form on the participant’s behalf after witnessing verbal consent.

Participants can optionally consent to receive trial newsletters, end-of-trial results, their allocated treatment group, and for potential longer-term follow-up via linkage to routinely collected clinical data.

### Confidentiality

All participant data will be collected and retained in accordance with the UK General Data Protection Regulation (UK GDPR), in conjunction with the Data Protection Act (DPA) 2018 and ICH GCP E6 R2.

Participants will be assigned a unique participant ID number and trial data will be reported anonymously in any publications. Personal identifiable data (PID) and contact details will be collected and stored securely in the study database and only used as required for the research (e.g., to send follow-up surveys/results).

Fields containing PID will be on separate eCRFs with access restricted. Mobile phone numbers will be shared with our SMS service provider, which is located in the United States, to send the follow-up texts; this will be outlined in the patient information sheet and on the participant consent documents.

PID will be stored securely for 10 years after the end of the study, after which all PID will be securely destroyed. The final dataset will be anonymised prior to being made available in the public domain for future research.

### Ancillary and post-trial care

After the end of trial participation, participants will continue to receive standard NHS care with no special arrangements made in relation to the trial.

If there is negligent harm during the clinical trial NHS Indemnity covers NHS staff, medical academic staff with honorary contracts, and those conducting the trial. NHS indemnity does not offer no-fault compensation and is unable to agree in advance to pay compensation for non-negligent harm.

### Trial registration

The trial is registered on ISRCTN (ISRCTN76873478) on 11^th^ October 2024 (^2^).

### Protocol and statistical analysis plan

The protocol is available via the ISRCTN registry (ISRCTN76873478). Current protocol version 5.0, 7th January 2025. The statistical analysis plan will be made publicly available, details provided on request.

### Data sharing

After the end of the trial, the anonymised research data and related outputs will be stored indefinitely in an open research repository hosted by University of Exeter. This will be described on the participant consent documents. Further details are included in the DMP, available on request.

## Author contributions

All individuals listed as authors of this protocol paper will have made substantial contributions to its conception or design, or acquisition, analysis, or interpretation of data for the work; drafted the work or revised it critically for important intellectual content; given final approval of the version to be published; and agree to be accountable for all aspects of the work. Specific contributions are outlined below:

- EC and FH; joint lead applicants and chief investigators, study design, preparation and drafting of protocol, preparation and drafting of manuscript.
- KJ; preparation and drafting of protocol, preparation and drafting of manuscript, overseeing study set-up and conduct.
- RL; preparation and drafting of manuscript, overseeing study conduct and acquisition of data.
- HT, AH, SC, OMW, DR study design, preparation and drafting of protocol, review of manuscript.
- PD: development of data capture/management systems, review of manuscript.
- JC, DA, EC, HC, CH, DL, MJR, DS, HEW, EG, JC; preparation of protocol, review of manuscript.

## Data Availability

All data produced in the present study are available upon reasonable request to the authors

## Acknowledgements

The study team acknowledges the support of all patient and public contributors to the trial, including the North Bristol Infection Science Patient Advisory Group, people who inject drugs (PWID) and patients with skin of colour who have contributed to the design and ongoing conduct of the trial. We thank Bristol Drugs Project for sharing their expertise in making research suitable for PWID and the UK Dermatology Clinical Trials Network for their ongoing support in trial design and promotion. We thank North Bristol NHS Trust for sponsoring the trial and providing their support, the Exeter Clinical Trials Unit, University of Exeter, for day-to-day trial coordination and data management, Modepharma Ltd for supply of the IMP and matched placebo, and the Centre for Healthcare Randomised Trials, University of Aberdeen, for the randomisation service. We also thank the members of the Trial Steering Committee (TSC) and Data Monitoring Committee (DMC) for their input and ongoing oversight of the trial. Finally, we thank Julie Gibbs for supporting the patient and public involvement (PPI) activities for the trial, and Heather McLachlan for her significant contributions to the set up and ongoing conduct of the trial.

## Appendix 1 – Risk assessment of short term side effects of dexamethasone

### Hyperglycaemia

We are including people with diabetes in this trial, due to their increased risk of cellulitis. However, dexamethasone can produce prolonged hyperglycaemia in people with diabetes, requiring insulin or other medications to control it. The absolute risk of severe hyperglycaemia with dexamethasone use appears low from recent trial data, however, given the potential seriousness of this risk, site-specific training and work instructions will be provided to sites covering hyperglycaemia, sick day rules, and advice on diabetes management. Participants with diabetes will be given additional patient information sheets to ensure they are fully informed prior to consent. These incorporate Diabetes UK guidance on sick day rules and how to manage high sugar levels. Cases of severe hyperglycaemia (ketoacidosis, hyperglycaemic hyperosmolar state or hyperglycaemia requiring new use of insulin) will be reported as serious adverse events in this trial and monitored by the trial oversight committees.

### Gastrointestinal (GI) toxicity/bleeding

The risk of GI bleeds is relatively low with corticosteroids; however, this risk may increase with other factors including peptic ulcer disease, use of non-steroidal anti-inflammatory drugs (NSAIDs) and older age. People with active gastric or duodenal ulceration will not be eligible for this trial due to this increased risk. As NSAIDs may be prescribed as part of standard practice to manage the pain of cellulitis and given the short course of corticosteroids being used and the fact that NSAIDs are not listed as a contraindication in the dexamethasone SmPC, co-administration is allowed for this trial. To mitigate any risk, sites are provided with a working instruction on assessing the risk of GI bleeding in patients. Clinicians are advised to consider prescribing a proton pump inhibitor if felt to be clinically indicated, based on the participant’s risk factors and recent NSAID usage. Cases of gastrointestinal bleeding will be reported as serious adverse events in this trial and monitored by the trial oversight committees.

### Psychosis

Psychosis is a recognised and serious complication of corticosteroid treatment, but it is related to dose and length of corticosteroid treatment.^32^ It is therefore likely to be a very rare event in this trial given the short course and low dose of dexamethasone being used. Cases of psychosis will be reported as serious adverse events in this trial and monitored by the trial oversight committees.

## Appendix 2 – Numerical rating scale of pain wording

Participants will receive the following wording either via SMS or spoken to them by a member of the trial team if SMS if data is being collected in person or on the phone.

“Please rate the level of pain you are currently experiencing due to your cellulitis. The scale is from 0-10. Where 0 means no pain at all and 10 means the worst pain you can imagine.”

